# The Disease Burden of Hereditary Pancreatitis on Children and Their Families: A Systematic Review

**DOI:** 10.1101/2025.10.10.25337728

**Authors:** Deki Choden Thinley, Fiona Campbell, David Campbell, Gurdeep Sagoo

**Affiliations:** School of Biomedical, Nutritional and Sport Sciences, Newcastle University, Newcastle Upon Tyne, UK; Population Health Sciences Institute, Newcastle University, Newcastle Upon Tyne, UK; Paediatric Gastroenterology, The Great North Children’s Hospital, Newcastle Upon Tyne Hospitals NHS Foundation Trust, Victoria Rd, Newcastle Upon Tyne, NE1 4LP

**Keywords:** Hereditary pancreatitis, disease burden, systematic review

## Abstract

**Objective:** To systematically review existing research evidence on the disease burden of hereditary pancreatitis (HP) in children, focusing on physical, psychological, social and financial consequences for families.

**Methods:** The review followed Synthesis Without Meta-analysis (SWiM) and PRSIMA guidelines. A structured search of PubMed, Google Scholar, and EMBASE was conducted. The PCC (Population, Concept, Context) framework guided study selection, targeting children (0-18 years) with HP and assessing physical, psychological, socioeconomic impacts. Data was extracted using a standardised form, and study quality was appraised using the Joanna Briggs Institute Checklist. Due to heterogeneity, a narrative synthesis was conducted using thematic analysis.

**Results:** Six studies met inclusion criteria. Children with HP presented chronic abdominal pain and hospitalisations, with median lifetime hospitalisations of 4 in chronic versus 1 in acute recurrent cases. Exocrine pancreatic insufficiency (EPI) were more common in chronic pancreatitis (CP) (33% vs 10%), and diabetes occurred in 8.7% versus 5.9%. Psychological distress was prevalent, with 19% showing clinically significant internalising behaviours and reduced quality of life. Up to 70% missed ≥1 school day per month, with greater pain-related social interference among females. High treatment costs with annual treatment exceeding $40,000 per patient.

**Conclusion:** HP significantly affects the physical and mental health of children and imposes financial and social strain on families. A structured, multidisciplinary approach to care including improved medical management, psychological support, and financial assistance is essential. Further research is required to develop targeted interventions aimed at managing psychological distress, reducing hospitalisations, and improving the quality of life of these children.

**What is already known:**

- Hereditary pancreatitis (HP) is a rare genetic disorder, associated with lifelong complications including abdominal pain, nausea and increased risk of pancreatic cancer.
- Evidence on wider psychosocial and financial impacts on affected children and their families has been fragmented and underreported.

**What this study adds:**

- This review shows that HP disrupts education, increases psychosocial distress, and imposes heavy financial burden on the healthcare system.

**How this study might affect research, practice or policy:**

- Multidisciplinary care models that include psychological and social support, alongside medical management, are urgently needed.
- Further research should focus on targeted interventions to reduced hospitalisations and improve quality of life in affected children.

## Introduction

Hereditary pancreatitis (HP) is a rare genetic disorder characterised by recurrent episodes of pancreatitis, which often begin in childhood. Most cases involve mutations in the PRSS1 gene, with others linked to SPINK1, CFTR, and CTRC. These mutations causes premature trypsin activation in the pancreas, leading to inflammation, fibrosis, and progression to chronic pancreatitis (CP)^1^. Symptoms often begin before age 10 and may include chronic pain, exocrine pancreatic insufficiency (EPI), and increased cancer risk^2^. The burden of HP extends beyond physical symptoms, encompassing psychological, social, and financial consequences^3^. Children commonly experience pain-related sleep disruption, missed school, social isolation and psychological distress such as anxiety or depression^4,5^. Despite this, psychological support is rarely standardised. Tools like the Paediatic Quality of Life Inventory (PedsQL)^6^ have been used to assess the psychological burden of HP on children. Families of affected children face substantial challenges, including frequent hospitalisations, complex medical management, and financial strain due to treatment costs^5,7^. Siblings may also experience emotional disruption^8^. Access to specialised care is often limited, particularly in low resource setting^5^. Current treatment focuses on symptoms management, including enzyme therapy, nutritional support^4^, and in severe cases, surgical options like total pancreatectomy with islet auto-transplantation (TPIAT)^9^. However, such procedure are not widely available and pose risks^10^. Although HP shares burden similarities with chronic conditions like cystic fibrosis^11^, inflammatory bowel disease^12^, and type 1 diabetes^13^, comprehensive support models are lacking^14,15,16^. This review systematically examines the physical, psychological, social, and financial burden of HP in children to inform clinical care and policy.

## Methods

### Study Design

This systematic review was conducted using the Synthesis Without Meta-analysis (SWiM)^17^ and reported under the Preferred Reporting Items for Systematic Reviews and Meta-Analyses 2020 guidelines^18^ to ensure a rigorous and transparent synthesis of findings.

### Eligibility Criteria

Inclusion criteria were defined using the PCC (Population, Concept, Context) framework^19^.

- Population (P): Children (aged 0-18 years) diagnosed with HP, including acute or chronic pancreatitis with hereditary causes and idiopathic origins.
- Concept (C): Disease burden, encompassing physical, psychological, social, and financial impacts on children and their families.
- Context (C): No restrictions on geographical location or publication period. Studies from various healthcare settings were included.

Eligible studies included observational, qualitative, and mixed methods designs published in English. Exclusions were studies focused only on adults, unrelated to pancreatitis (even if the term “pancreatitis” was mentioned without direct hereditary relevance), case reports, reviews, and opinions pieces without burden-related outcomes.

### Search Strategy

Comprehensive searches were conducted in PubMed, Google Scholar, and EMBASE using MeSH terms, Boolean operators, and free-text keywords. Example search strings included:

- (“hereditary pancreatitis” OR “HP”) AND (“disease burden” OR “impact”) AND (“children” OR “paediatric” OR “youth”) AND (“families” OR “caregivers” OR “parents”)
- (“hereditary pancreatitis” OR “HP”) AND (“long-term effects” OR “progression”) AND (“children” OR “paediatrics”) AND (“family burden” OR “caregivers experience”)

### Study Selection

Screening was conducted in two stages: (1) titles and abstracts were assessed for relevance, and (2) full-text articles were reviewed for eligibility and reasons for exclusion recorded. A pilot test on a subset of studies refined inclusion and exclusion criteria. EndNote was used to manage references and track screening decisions. The selection process is presented in the PRISMA flow diagram (Figure 1).

**Figure 1:**
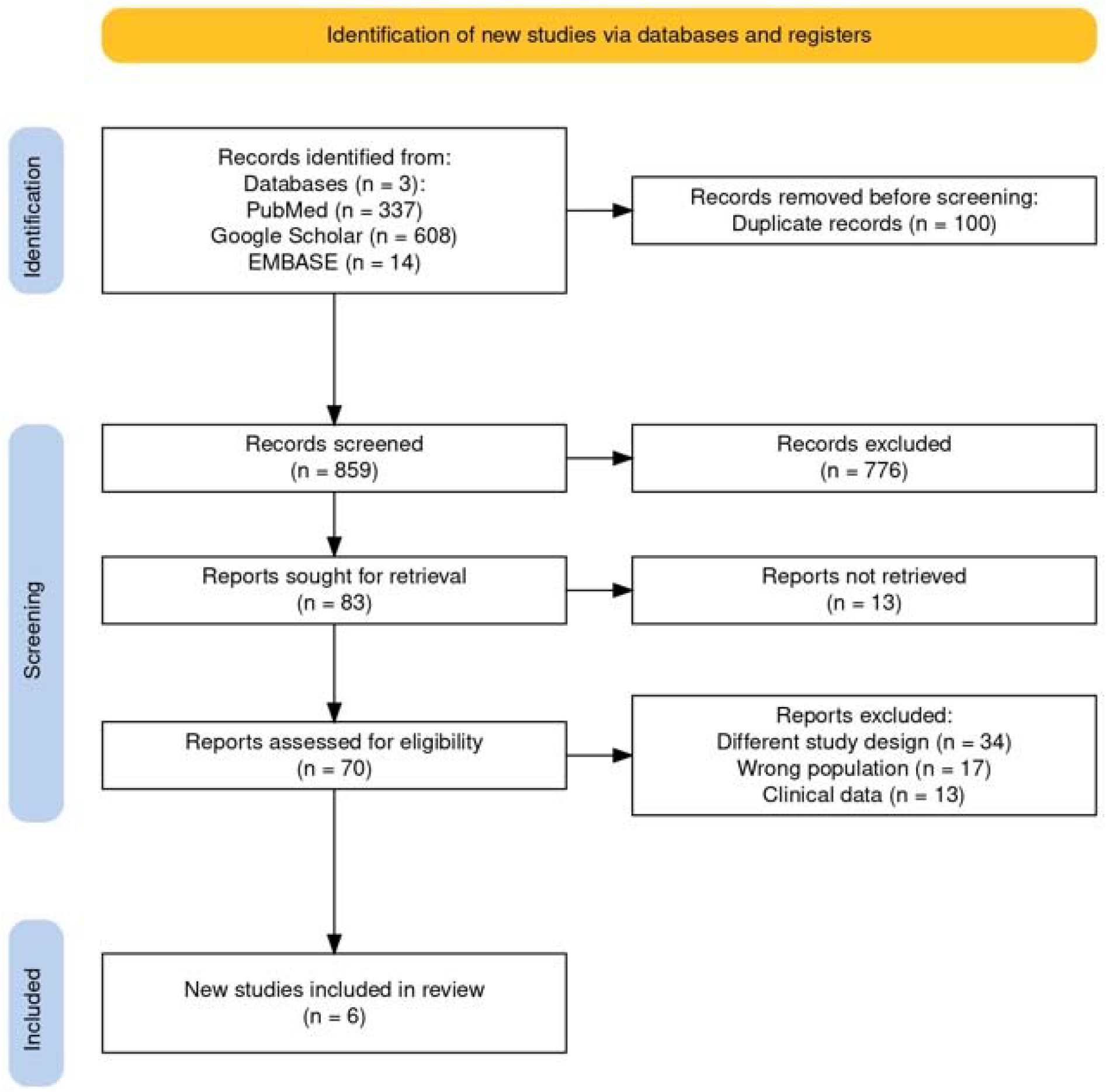
Illustration of study selection process using the PRISMA flowchart. This flowchart depicts the stages of study selection, starting from the initial identification of studies, through the screening process, full-text review, and final inclusion of studies that met the inclusion criteria for the synthesis.

### Quality Assessment

The quality of the included studies was evaluated using the Joanna Briggs Institute (JBI) Critical Appraisal Checklist for Analytical Cross-Sectional Studies^20^ (Figure 2).

**Figure 2:**
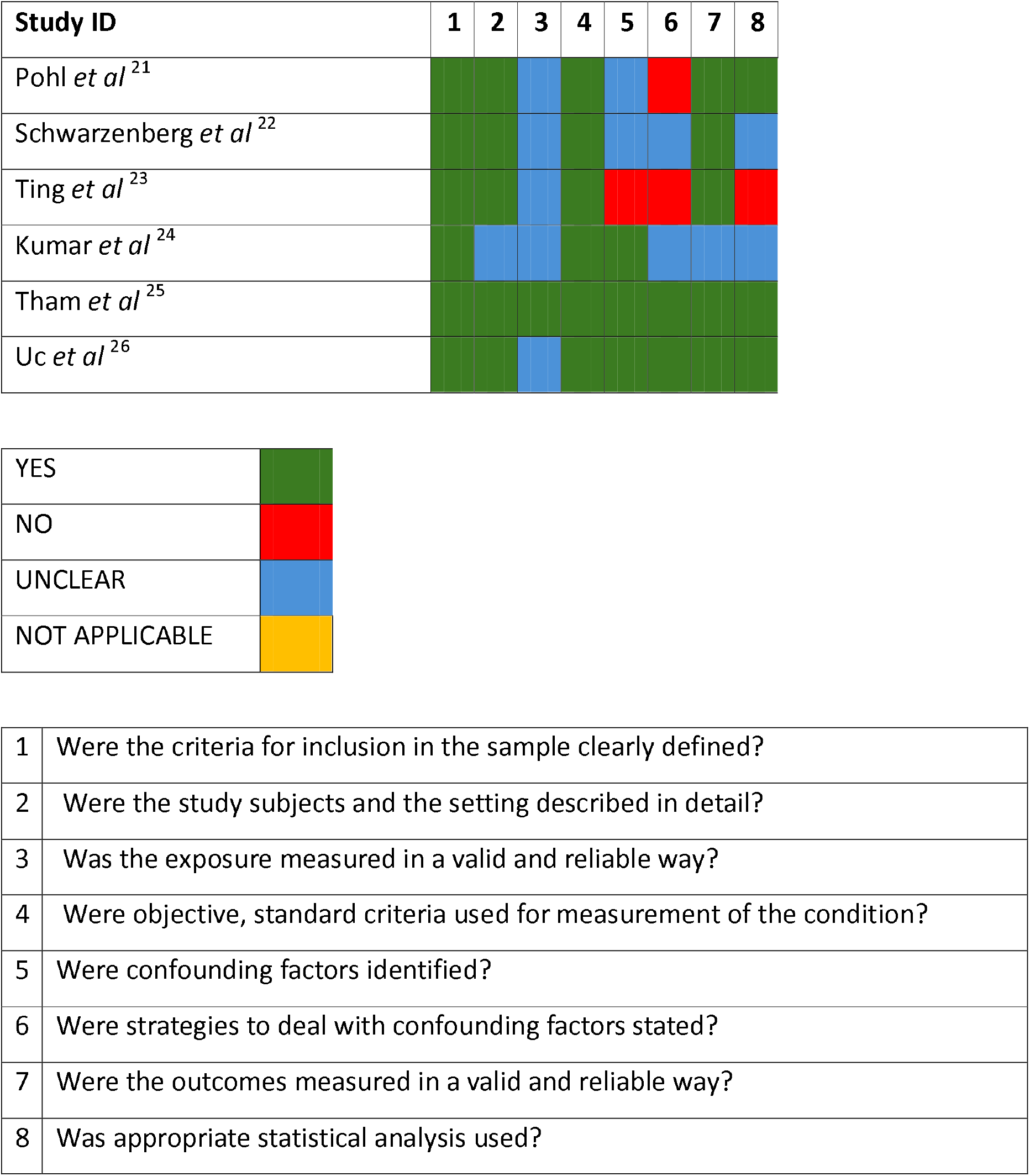
Assessment of study quality using the JBI Critical Appraisal for Analytical Cross-Sectional Studies for the Included Studies. This figure displays the quality evaluation of the included studies, based on key criteria such as clarity in sample inclusion, valid measurement of exposures and outcomes, identification and control of confounding factors, and use of appropriate statistical analysis.

### Data Extraction

A standardised data extraction form was used to collect study details (author(s), year, country, study location(s), design, sample size), populations characteristics (age range, standardised for comparison across studies), outcome measures, and key findings (Table 1).

**Table 1:**
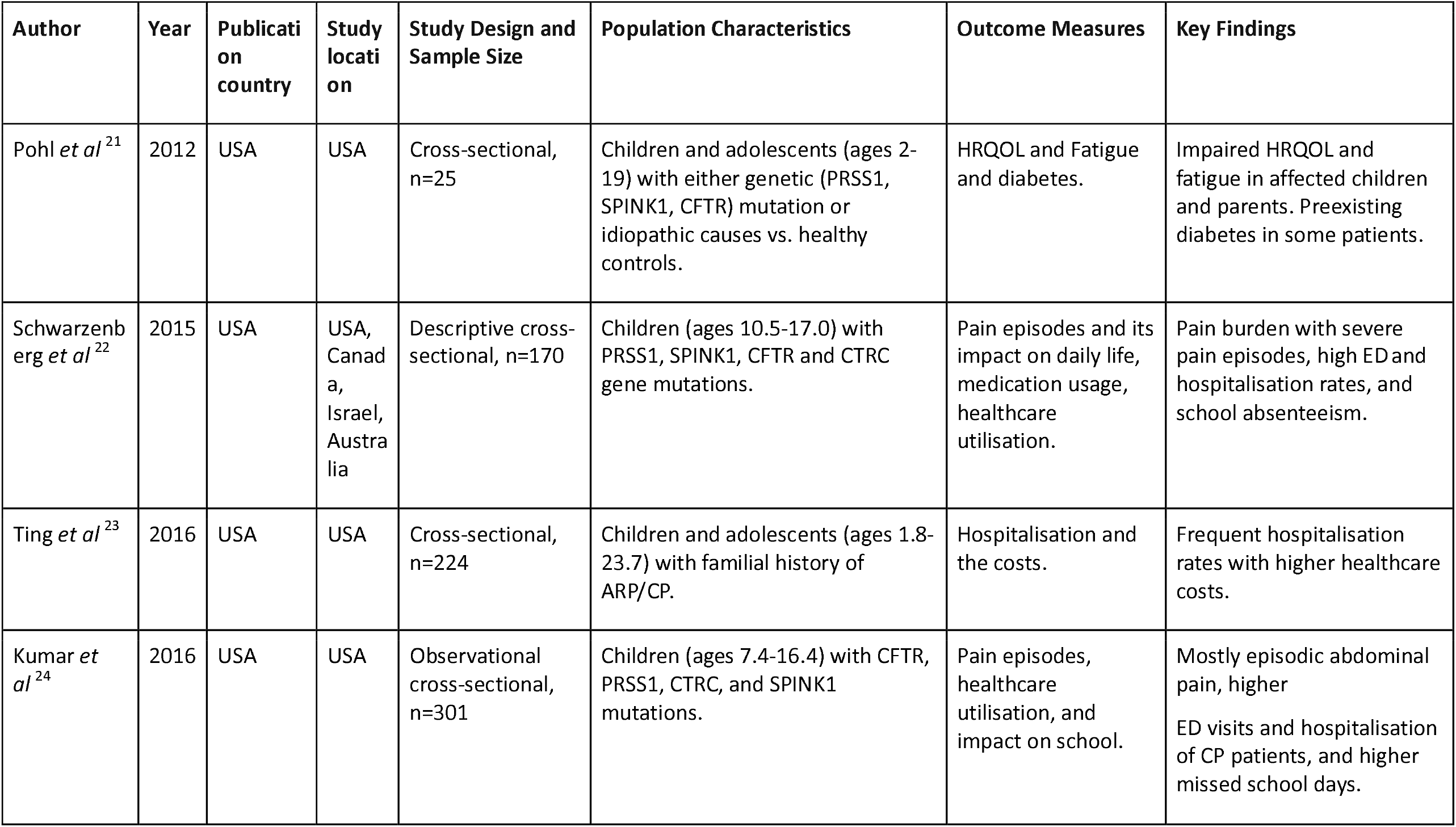
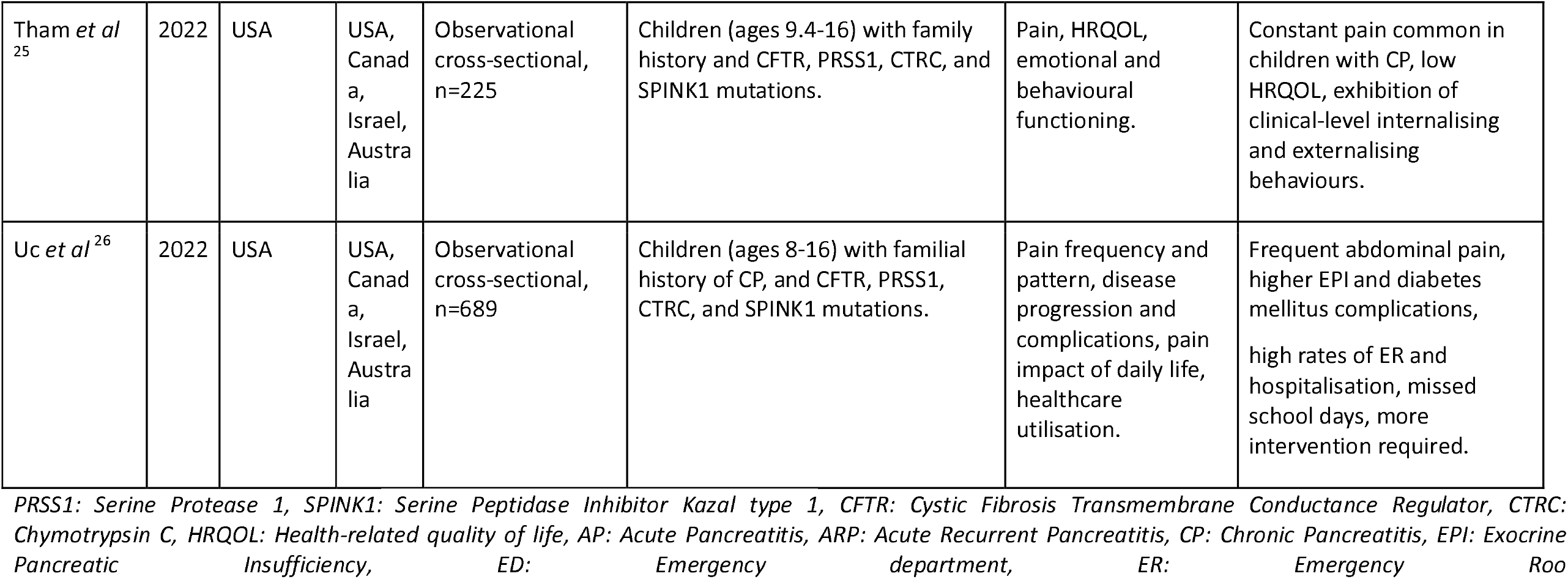
Study Characteristics. This table provides an overview of the key characteristics of the studies included in the review, including study design, sample size, population, and other relevant factors.

| Author | Year | Publication country | Study location | Study Design and Sample Size | Population Characteristics | Outcome Measures | Key Findings |
| --- | --- | --- | --- | --- | --- | --- | --- |
| Pohl *et al*<sup>21</sup> | 2012 | USA | USA | Cross-sectional, n=25 | Children and adolescents (ages 2-19) with either genetic (PRSS1, SPINK1, CFTR) mutation or idiopathic causes vs. healthy controls. | HRQOL and Fatigue and diabetes. | Impaired HRQOL and fatigue in affected children and parents. Preexisting diabetes in some patients. |
| Schwarzenberg *et al*<sup>22</sup> | 2015 | USA | USA, Canada, Israel, Australia | Descriptive cross-sectional, n=170 | Children (ages 10.5-17.0) with PRSS1, SPINK1, CFTR and CTRC gene mutations. | Pain episodes and its impact on daily life, medication usage, healthcare utilisation. | Pain burden with severe pain episodes, high ED and hospitalisation rates, and school absenteeism. |
| Ting *et al*<sup>23</sup> | 2016 | USA | USA | Cross-sectional, n=224 | Children and adolescents (ages 1.8-23.7) with familial history of ARP/CP. | Hospitalisation and the costs. | Frequent hospitalisation rates with higher healthcare costs. |
| Kumar *et al*<sup>24</sup> | 2016 | USA | USA | Observational cross-sectional, n=301 | Children (ages 7.4-16.4) with CFTR, PRSS1, CTRC, and SPINK1 mutations. | Pain episodes, healthcare utilisation, and impact on school. | Mostly episodic abdominal pain, higher ED visits and hospitalisation of CP patients, and higher missed school days. |
| Tham *et al*<sup>25</sup> | 2022 | USA | USA, Canada, Israel, Australia | Observational cross-sectional, n=225 | Children (ages 9.4-16) with family history and CFTR, PRSS1, CTRC, and SPINK1 mutations. | Pain, HRQOL, emotional and behavioural functioning. | Constant pain common in children with CP, low HRQOL, exhibition of clinical-level internalising and externalising behaviours. |
| Uc *et al*<sup>26</sup> | 2022 | USA | USA, Canada, Israel, Australia | Observational cross-sectional, n=689 | Children (ages 8-16) with familial history of CP, and CFTR, PRSS1, CTRC, and SPINK1 mutations. | Pain frequency and pattern, disease progression and complications, pain impact of daily life, healthcare utilisation. | Frequent abdominal pain, higher EPI and diabetes mellitus complications, high rates of ER and hospitalisation, missed school days, more intervention required. |

PRSS1: Serine Protease 1, SPINK1: Serine Peptidase Inhibitor Kazal type 1, CFTR: Cystic Fibrosis Transmembrane Conductance Regulator, CTRC: Chymotrypsin C, HRQOL: Health-related quality of life, AP: Acute Pancreatitis, ARP: Acute Recurrent Pancreatitis, CP: Chronic Pancreatitis, EPI: Exocrine Pancreatic Insufficiency, ED: Emergency department, ER: Emergency Room

### Data Synthesis

Due to heterogenicity, data were narratively synthesised using the SWiM (Table 2). A thematic analysis grouped findings into four domains: (1) physical health burden (pain, hospitalisations, diabetes, EPI risk), (2) psychological burden (emotional and behavioural impact), (3) social (school absences) and (4) financial burden (financial strain).

**Table 2:**
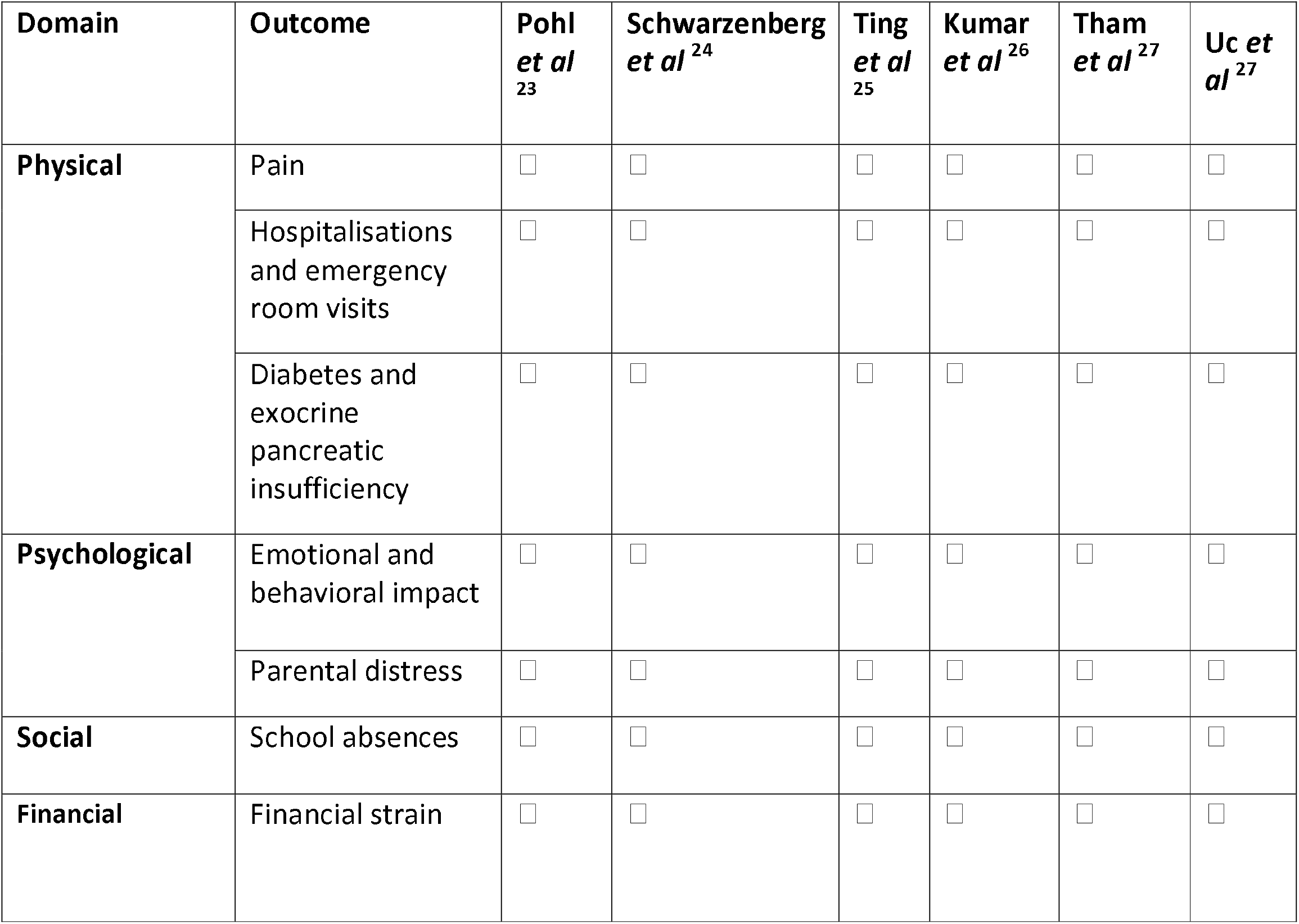
Summary of Burden outcomes Across Studies. This table shows the presence (□)or absence (□) of outcomes grouped by burden domain across the six included studies.

### Ethical Considerations

As this review used published data, ethical approval was not required; ethical guidelines for systemic reviews were followed with appropriate source attribution.

### Use of AI tools

AI-based tool, ChatGPT (OpenAI) was employed to assist in summarising complex concepts and improvising cohesiveness throughout the manuscript. All AI-generated content was critically reviewed and edited by the author, who is fully responsible from the accuracy and integrity of the final work.

## Results

The database search identified 959 records. After removing duplicates, 859 titles and abstracts were screened, with 776 excluded. Seventy full texts were assessed, and six studies met the inclusion criteria for synthesis. The selection process is illustrated in the PRISMA flow diagram (Figure 1).

### Study Characteristics

The six included studies, all cross-sectional, varied in populations, settings, and findings. Sample sizes ranged from 38 to 689, with a total of 1,790 children diagnosed with HP. Ages were standardised for comparability, with a minimum of two years (Table 1).

Most studies used data from the INSPIRE registry^22,23,25,26^, with one contributing additional cases^24^ and another based on medical records from a single centre^21^. All were published in the United States, through several incorporated international cohorts from Canada, Isreal, and Australia^22,25,26^.

US-only studies ^21,23,24^ provided detailed data relevant to the American healthcare contact but limited generalisability. Multi-country studies^22,25,26^ improved diversity and comparative insights, though they faced logistical and funding challenges. Together, these studies offer complementary perspectives, highlighting both national trends and global variations in disease burden and healthcare outcomes.

### Synthesis of Findings

This review explored the burden of HP on children and their families across three domains: physical health, psychological impact, social and financial consequences. A narrative synthesis using the SWiM framework highlighted the following (Table 2).

#### Physical Health Burden

Abdominal pain was the predominant symptom in both acute recurrent pancreatitis (ARP) and CP patients across all studies^21-26^. Children with pancreatitis experiences significantly lower health-related quality of life (HRQOL), particularly in general health and bodily pain, with persistent pain associated with the poorest outcomes^25^. Fatigue was common and strongly linked to lower psychical HRQOL^21^. Healthcare utilisation was substantial. Among 57 CP patients, only one had no hospitalisations or emergency department (ED) visits; median ED visits were 3.0 (IQR 1-5) and hospitalisation 2.0 (IQR 1-3). Constant pain patterns were associated with more ED visits than episodic patterns (median 3.0 vs 1.0, P=0.01)^22^.

CP patients demonstrated greater healthcare utilisation than ARP patients, including more ED or emergency room (ER) visits over their lifetime [(median [IQR] 4.5 [1–10] vs. 2 [1–4]; 95% CI, 1.4–3.6; P < 0.001)^24^, (median, 6-7 vs. 2)^26^]. Hospitalisations were also higher in CP patients, with greater median lifetime hospitalisations (median [IQR] 4 [1–7] vs. 1 [1–4]; 95% CI, 1.0–3.0; P < 0.001)^24^ and higher past-year rates (median, 1 vs 0) compared with ARP patients^26^.

EPI was more common in CP than ARP (33% vs 10%, P<0.001)^21^, while diabetes mellitus prevalence was slightly higher in CP (8.7% vs 5.9%) but not statistically significant^26^. Among CP patients, three required insulin therapy, and impaired glycaemic control often preceded severe pain onset^21^. This factor did not significantly modify the association between HP and subsequent risk estimates.

#### Psychological Burden

Children with ARP and CP showed elevated internalising behaviours (anxiety or depression) and total behavioral difficulties relative to normative expectations, reflecting trends observed in the broader sample studies^25^, in which 19.3% showed clinically significant internalising behaviours, 5.2% exhibited clinical levels of externalising behaviours (oppositional and aggressive behaviours), and 11.4% had high total problems scores. Children with long-standing pancreatitis and their parents reported notably lower HRQOL and greater fatigue compared to healthy children, with moderate agreement between child and parent reports^21^.

#### Social Burden

In CP cases, 70% of patients missed ≥1 day of school per month, and 34% missed ≥3 days per month^22^. Median absences in patients with constant pain were 8 days/month (∼2 days/week), corresponding to approximately 40% of school lost and 60% attendance. In contrast, children with episodic pain had a median of 1.5 days/month (∼0.4 days/week) missed, equating to ∼8% of school lost and ∼92 attendance. Another cohort reported a median of 2 days/month missed in CP (∼0.5 days/week; ∼10 lost; 90% attendance)^24^. In addition, females with CP experienced greater pain-related interference with school and social activities compared with males, likely reflecting the higher proportion of female with CP in the cohort^26^.

#### Financial Burden

The average annual direct cost per patient was estimated at $40,589, encompassing both outpatient medications and inpatient treatments. Inpatient medication costs ranged from $614 to $4,114, depending on treatment type. Total estimated costs for managing HP in the United States ranged from $60 million to $70 million annually. Hospitalisations averaged 2.3 per patient per year, with a mean cost of $38,755 per hospitalisation. Surgical procedures added approximately $42,951 per patient per year. Notably, no studies included in this review reported secondary or indirect costs, such as lost parental income or travel expenses^23^.

#### Quality Appraisal Findings

Compliance to the JBI Critical Appraisal for Analytical Cross-Sectional Studies^20^ is summarised in Figure 2. The included studies reported common limitations in unclear identification and measurement of potential confounding factors^21-23^, as well as the failure to implement strategies to address these confounders^21-24^. Additionally, several studies^21-24,26^ failed to provide a detailed discussion of the validity and reliability of the exposure measured and lacked formal statistical analyses of the data^22,23,24^. One study^24^ did not provide a clear and detailed description of the subjects, setting and outcome measures.

## Discussion

This systematic review assessed the burden of HP on children and their families, examining physical, psychological, and social-financial impacts. Persistent pain, frequent hospitalisations, and long-term complications such as diabetes, significantly reduce health-related quality of life. Psychological distress affects both children and parents, cascading into social and educational challenges. Financial strain further compounds this burden, underscoring the need for targeted healthcare strategies. Previous studies have detailed the physical effects of HP, including inflammation and pancreatic necrosis^27^. Liu *et al*^28^ highlighted rapid progression in children with PRSS1 mutations, with a median transition to CP within 2.25 years, accompanied by persistent pain. This review confirms as pain as a dominant, enduring challenge requiring ongoing medical management^23,25^.

The psychological burden of HP, underexplored in prior reviews^5^, includes emotional distress and behavioural challenges related to recurrent pain. The unpredictable nature of pain often contributes to emotional dysregulation and maladaptive coping, such as medications overuse, reflecting patterns seen in other chronic paediatric conditions^29^. Parental distress is significant, with anxiety often intensified after genetic testing^30^. Parents, particularly mothers, report guilt and fear over disease transmission and caregiving burdens, which can strain relationships and lead to social isolation^31^. Socially, frequent hospitalisations causes school absences, hampering academic progress and peer relationships^32^. Children with chronic illnesses are more likely to underperform academically and experience social isolation due to stigma^33^ and physical limitations^34^, contributing to loneliness^35^ and depression^36^. Although there are no direct measurements of how education days lost to HP effect subsequent lifetime potential earnings, it can only be deduced that days lost affect both the child in the long run and the family immediately, with an adverse financial consequence. Families face significant direct costs related to hospital visits, diagnostics, enzyme therapy, and surgeries. Shelton *et al*^37^ noted 43% of families perceive medical expenses as burdensome, with data supplementing HP-related healthcare costs exceeding $2 billion annually^38^ in the US alone. Among treatment options, TPIAT has proven effective in improving physical and emotional quality of life in paediatric patients^39^. However, the procedure’s high upfront costs—including surgery, hospitalisation, and follow-up care— place significant pressure on families and healthcare systems^22^. Despite the initial expenses, the NHS^40^ suggests that TPIAT may be cost-effective long term, as it reduces reliance on pain management, recurrent hospitalisations, and other medical interventions compared to conservative management.

The quality of the included studies critically influenced the strength and reliability of the conclusions drawn from this systematic review. There were significant methodological concerns, particularly regarding confounding factors and measurement validity. The failure to define, measure, or control for confounders increased the risk of biased associations, complicating the determination of causal relationships between variables. Additionally, the lack of discussion on the validity and reliability of exposure and outcome measures introduced uncertainty regarding the accuracy and consistency of the reported findings. Without robust statistical analyses, the studies lacked the rigour needed to produce precise and generalisable results. Collectively, these limitations reduced the overall confidence in the findings and emphasise the need for larger, more rigorously designed studies in future research.

This review advanced the understanding of how the disease burden of paediatric HP affects children and families by integrating both quantitative healthcare data and qualitative family impact measures, a key gap in prior research. By synthesising these perspectives, it highlighted the broader effects on families, moving beyond individual patient outcomes. Notably, it incorporated children’s own voices, a rarely explored dimension in paediatric pancreatitis research. Furthermore, this review provided a comprehensive estimate of both direct and indirect economic costs associated with the condition. However, several limitations involved the heterogeneity of study designs and outcome measures precluded meta-analysis, necessitating a narrative synthesis. This approach limited the strength of clinical recommendations. Some included studies relied on small, single-centre samples limiting the generalisability of the findings. Furthermore, majority of the included studies focused on tertiary care centres, potentially excluding milder cases and introducing bias in the disease representation.

Although this review primarily focused on the direct impacts of pancreatitis, long-term follow-up data beyond adolescence remains sparse. Future studies should prioritise larger, multi-centre studies to capture a more comprehensive range of long-term psychosocial outcomes for individuals who survive into adulthood, particularly regarding academic achievement, career trajectories, and family life. Economic data in the included studies were primarily derived from developed healthcare systems, limiting the applicability of the findings to low-income or non-western healthcare contexts. Moreover, the exclusion of unpublished studies and non-English articles introduced the possibility of publication bias, which may distort the overall conclusions by under-representing certain findings or perspectives.

The findings from this review emphasise the need for more research on the early identification of HP and the improvement of genetic testing and awareness. Early detection through genetic screening can facilitate better disease management and interventions, potentially reducing the severity of the condition. Raising public and clinical awareness of HP is also essential for timely diagnosis and intervention, minimising the long-term impact on affected children and their families. A multidisciplinary approach to managing paediatric HP is essential, integrating medical, psychological, and social support. Clinicians should prioritise mental health care by incorporating routine screenings for anxiety and depression and fostering peer support networks to enhance the affected children’s overall well-being. Healthcare providers must train medical staff to educate, and support affected families, ensuring they have the necessary skills to deliver comprehensive care. Policy reforms are urgently needed to improve access to care for families affected by HP. In private insurance-based systems, this includes expanding insurance coverage for the long-term treatment of HP. In publicly funded systems, efforts^15^ should ensure adequate resources and equitable access. Mental health support must be integrated into care plans to address the psychological burden on patients and families. There is also a need for studies on the effectiveness of interventions aimed at reducing the disease burden, including psychosocial support programmes and educational accommodations for affected children.

## Conclusion

In conclusion, HP imposes a significant, multi-dimensional disease burden on both children and their families. The physical burden, including chronic pain and increased risk of pancreatic cancer, severely impacts the child’s quality of life. Alongside these physical challenges, there is a considerable psychological burden, with emotional and behavioural effects on the children, and significant distress experienced by parents. Additionally, the financial strain from treatment and medical care further exacerbates these challenges. These burdens not only affect the child’s well-being but also compromise the stability and resources of the family unit. Addressing these challenges requires a comprehensive, multidisciplinary approach that combines medical management, psychological support, and financial assistance to ease the strain on families. Future research should focus on developing interventions that can alleviate these burdens and improve the quality of life for both affected children and their families.

## Data Availability

All data produced in the present study are available upon reasonable request to the authors

## Contributors

Miss Deki Choden Thinley conducted the literature search, screened and analysed the studies and drafted the manuscript. Dr Gurdeep Sagoo assisted in drafting the initial version of the manuscript. Dr Fiona Campbell and Dr David Campbell revised the manuscript, provided critical feedback, and contributed to the editing of the final version. All authors read and approved the final manuscript.

## Licence statement

I, Deki Choden Thinley, the Submitting Author has the right to grant and does grant on behalf of all authors of the Work (as defined in the below author licence), an exclusive licence and/or a non-exclusive licence for contributions from authors who are: i) UK Crown employees; ii) where BMJ has agreed a CC-BY licence shall apply, and/or iii) in accordance with the terms applicable for US Federal Government officers or employees acting as part of their official duties; on a worldwide, perpetual, irrevocable, royalty-free basis to BMJ Publishing Group Ltd (“BMJ”) its licensees and where the relevant Journal is co-owned by BMJ to the co-owners of the Journal, to publish the Work in Frontline Gastroenterology and any other BMJ products and to exploit all rights, as set out in our licence.

The Submitting Author accepts and understands that any supply made under these terms is made by BMJ to the Submitting Author unless you are acting as an employee on behalf of your employer or a postgraduate student of an affiliated institution which is paying any applicable article publishing charge (“APC”) for Open Access articles. Where the Submitting Author wishes to make the Work available on an Open Access basis (and intends to pay the relevant APC), the terms of reuse of such Open Access shall be governed by a Creative Commons licence – details of these licences and which <u>Creative Commons</u> licence will apply to this Work are set out in our licence referred to above.

